# A Machine Learning Analysis of The Bead Maze Hand Function Test for Predicting Manual Dexterity in Children

**DOI:** 10.64898/2026.01.25.26344808

**Authors:** Aadit Narayanan, Komal K. Kukkar, Pranav J. Parikh

## Abstract

Comprehending how forces are applied to an object during manipulation can help provide important insights into the quality of behavior in daily tasks. We have developed the Bead Maze Hand Function test to objectively measure the quality of hand function in children. This test aims to measure how well an individual performs the activity by integrating measures of time and force control. The main objective of this study was to examine associations between a common clinical measure, the Box and Block Score (BBS), and variables on the Bead Maze Hand Function test that were either time-based or force-based. The sample was composed of neurotypical developing adolescent participants (N=23). We found that the time (duration) on the double curve wire (most complex) was the best predictor of BBT. Furthermore, we found that force-based variables were weak predictors of the clinical, time-based BBS. These findings support the integration of time and force-based metrics to holistically quantify the quality of motor behavior. Linking these metrics into a unified score may serve as a better way to analyze adolescent motor behavior and predict future motor or neurodegenerative conditions.

## I. INTRODUCTION

To effectively grasp and manipulate an object, a child needs to be able to control the amount of force production they use [1]. Force control is a sensorimotor behavior that develops throughout adolescence [10,11]. There are immediate consequences when the development of the ability to control forces is impaired [2,5]. For example, the potential inability to control forces while writing may lead to poor handwriting [5]. Moreover, lackluster force control in grasping and holding objects may result in events like accidental spills, drops, or the inability to complete mundane tasks [1,7]. By understanding the forces applied to an object during manipulation, these insights can reveal the relationship between force application and the quality of behavior [1,3,4,6]. Typical clinical hand function tests rely on subjective observation of the hand behavior and/or time-based measures but do not examine force control, which may neglect capturing important sensorimotor aspects like smoothness, force scaling, and trial variability [5,18]. Therefore, the current clinical tests may be less responsive to functional changes in hand function post-intervention(s) [5,18]. Based on previous results from laboratory-based studies of force control [1,3,4,10], we have developed an objective measure of the quality of hand function in children, the Bead Maze Hand Function Test (BMHFT). The BMHFT quantifies not only how quickly a task is accomplished (i.e., provides a time-based measure) but also how well the individual performs the activity by integrating measures of time and force control. During the test, children use their digits to slide a small bead over smooth wires of different shapes (straight, single-curve, and double-curve) that require small changes in movement direction, while force sensors measure the forces applied to the wire.

The main objective of this study was to determine which BMHFT measure best predicts manual performance on a widely used test of hand function, the Box and Block Test (BBT), in typically developing children. The BBT is scored by counting the number of blocks carried over the partition from one compartment to another during a one-minute trial [17]. We used various types of machine learning models to safeguard against common issues that arise from the high-feature dataset, like high correlation between variables, noise, and overfitting. Our previous work showed that the BMHFT measures obtained on the wire with the highest complexity (double-curve) were a more relevant measure of the quality of behavior [23]. We hypothesized that the time-based BMHFT variable on the wire with the highest complexity (double-curve) would be the strongest predictor of BBT. Due to poor correlation between the forces exerted on the object during object manipulation and the task completion time, we hypothesized that force-based variables obtained on the BMHFT would be weaker predictors of BBT. This study aims to extend our previous work by incorporating predictive modeling to identify important features in potentially non-linear relationships while also minimizing multicollinearity, with the end goal of providing novel insights into which experimental metrics show up in standard clinical measures.

## II. METHODS

The study included twenty-three neurotypical children (9 girls; age range = 4-16, M = 8.56, SD = 3.45) who were recruited locally by word of mouth and flyer over the course of a year. Older children were included because finger force control continues to develop throughout adolescence [10]. Participants had normal or corrected-to-normal vision, no history of developmental disorders, no neurological abnormalities, no musculoskeletal disorders, no history of hand/wrist injuries/surgeries, and attended a mainstream school as reported by a parent. Verbal assent was obtained from all participants, and informed written consent was obtained from the participants’ parents. All participants had no prior experience with the apparatus. The study was approved by the University of Houston Institutional Review Board.

From these participants, we analyzed data from both hands, yielding 46 observations. Data from both hands was analyzed independently, since lateralized dominance (handedness) had not developed for the majority of the participant sample based on age range. This methodological approach is appropriate for analyzing performance in pediatric populations during object manipulation, while acknowledging the possibility of residual within-subject correlations.

The Bead Maze apparatus used for the Bead Maze Hand Function Test (**Fig. 1**) has three force transducers (Nano 25, ATI Industrial Automation; 1,000 Hz) attached to a bottom plate and mounted on a wooden base. Straight and curved polished metal wires were attached through a top plate. This arrangement allowed us to monitor forces applied to the wire along the horizontal (x and y; **Fig. 2**) and vertical (z) axes. Note that the sensors are installed in the base of the device, and there are no sensors within the bead. Each wire is tightly and directly attached to the sensor to allow for continuous recording of forces as the bead is moved along the wire. Any slight pushing, tilting, or rotating of the bead, which may happen while moving the bead over the wire, will still transmit forces onto the wire and is thus measured by the attached sensor. Sensor calibration was verified prior to each session using static loads. Our apparatus uses three different colored wires that increase in complexity based on their shape (straight, single-curve, double-curve) to analyze motor skills at varied difficulties. This was to allow for effective assessments of motor quality both before and after intervention in all populations. Additionally, the varying and increasing difficulty among the wires prevents the test from reaching a ceiling effect, which allows for a more accurate analysis. The moving beads (MVBs; **Fig. 1**) that slide along the wire have an outside diameter of 18 mm, a lumen diameter of 6.5 mm, and are textured to improve grip and minimize slippage. Additionally, each wire also has a stationary bead (STB) affixed to it.

**Fig. 1.**
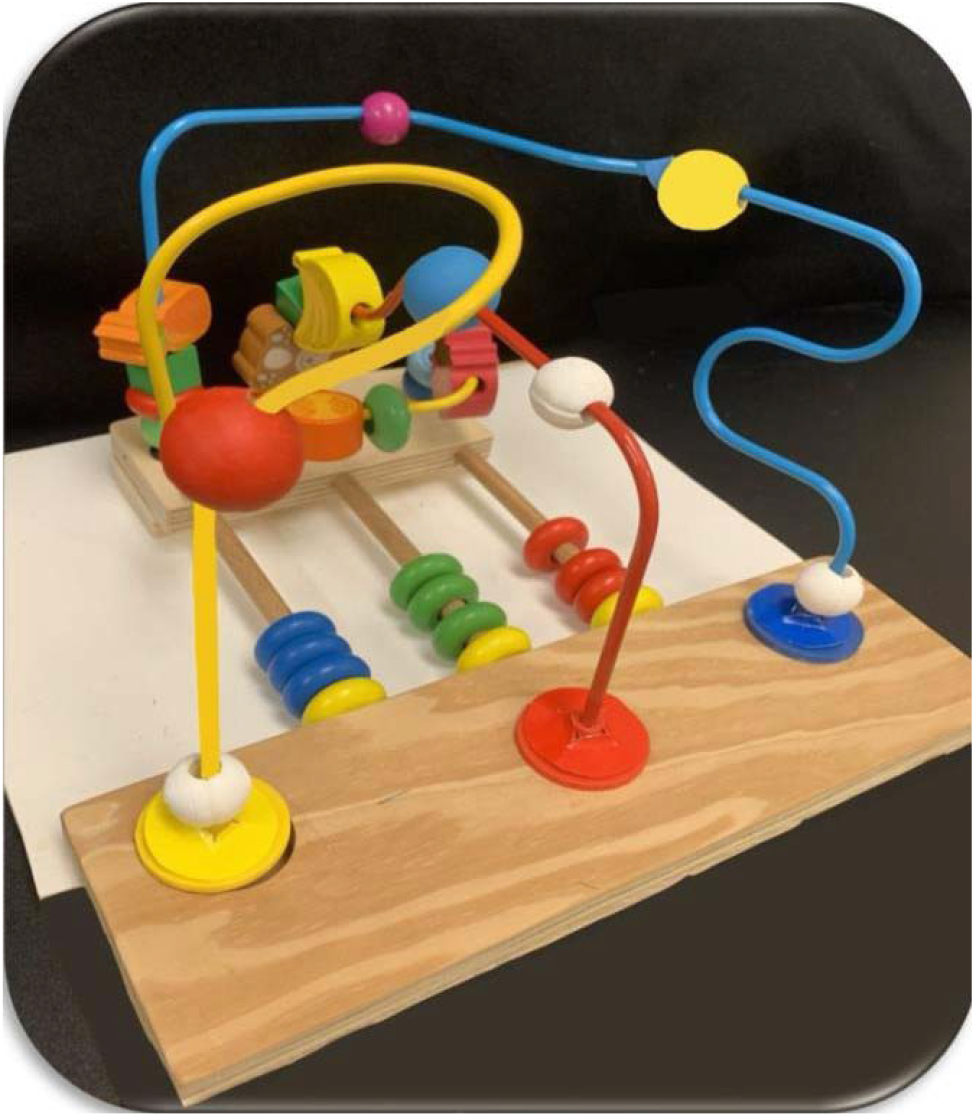
Illustration of Bead Maze Hand Function Test (BMHFT). Wires from left to right: Straight, Single Curve, Double Curve

**Fig. 2.**
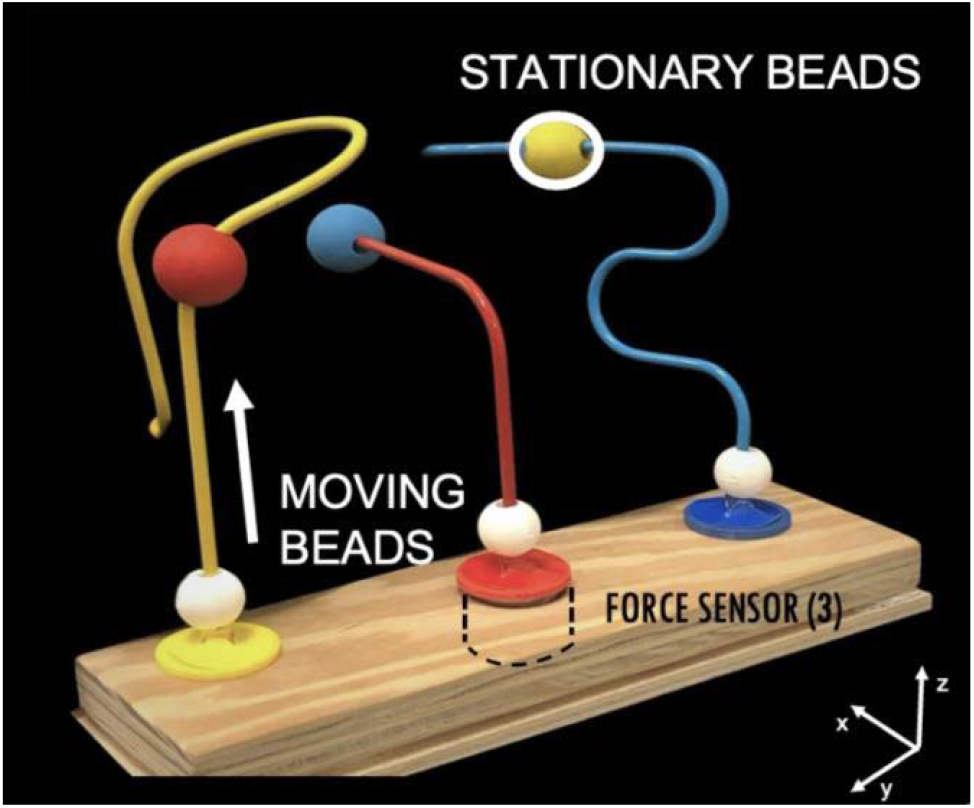
Bead Maze Hand Function Test (BMHFT) apparatus with integrated force sensors in the base enabling tri-axial force measurement.

### A. Bead Maze Hand Function Test (BMHFT)

For the test, participants sat on a chair and rested their hands on their lap. The Bead Maze device was positioned about 10 cm from the edge of the table. From there, participants were instructed to grasp the MVB, lift one, and move it along the wire until it “bumped” the STB at the end. Once the MVB contacted the STB, participants were instructed to release the grasp and return their hand to their lap. A demonstration of how the task should be performed was done by the experimenters, followed by one practice trial per wire for the participants. Participants then performed the task five times per wire at a self-chosen “comfortable” speed (blocked for each hand). The wire order was randomized and counterbalanced. Once the task had started, no instructions on how to perform the task, grip the bead, or touch the bead to the wire were provided. Instructions were intentionally minimized to preserve the naturalistic and playful nature of the task, as standardization of precision-speed constraints would be impractical for future use in younger or clinical populations.

### B. Data Analysis and Statistics

Data was acquired through a custom program in LabVIEW software (National Instruments) and analyzed using a custom-written script in Python 3.11. The initial contact with the MVB was defined as the time point at which the force produced in the x, y, or z direction was above a mean ±2 SD of the baseline for 10 ms. The end of the trial was defined by a stereotypical change in force caused by contact of the MVB with the STB during a phase just before forces returned to baseline (i.e., grasp release; Fig. 3–5). Trial end thresholds included +0.1 N z-force for Wire 1, +0.2 N y-force for Wire 2, and −0.2 N y-force for Wire 3. For each trial, the script identified the initial contact time and the end of the trial time (Fig. 3–5). For the reliability study, two independent researchers analyzed the outputs using the same custom-written script on separate computers. The researchers visually checked the times from the script in relation to the force traces for accuracy. In case of errors, the time values were adjusted manually. The absolute value of the force signals from contact to end were summed to the total force for each trial.

**Fig. 3.**
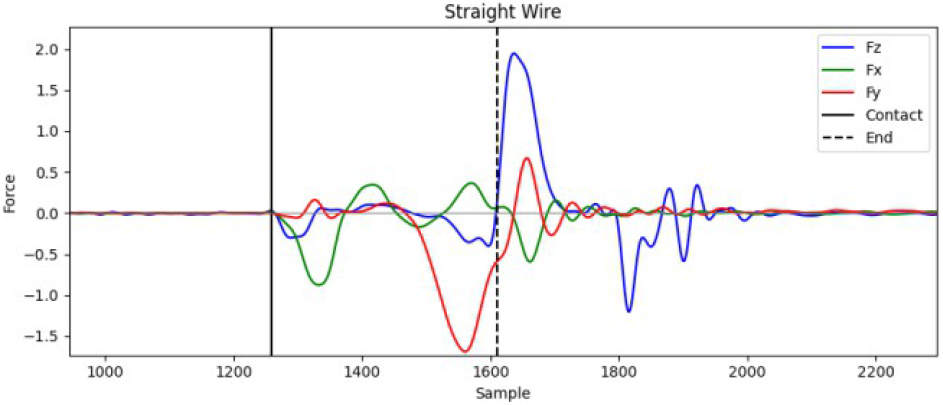
Raw force–time trace for the straight wire condition

**Fig. 4.**
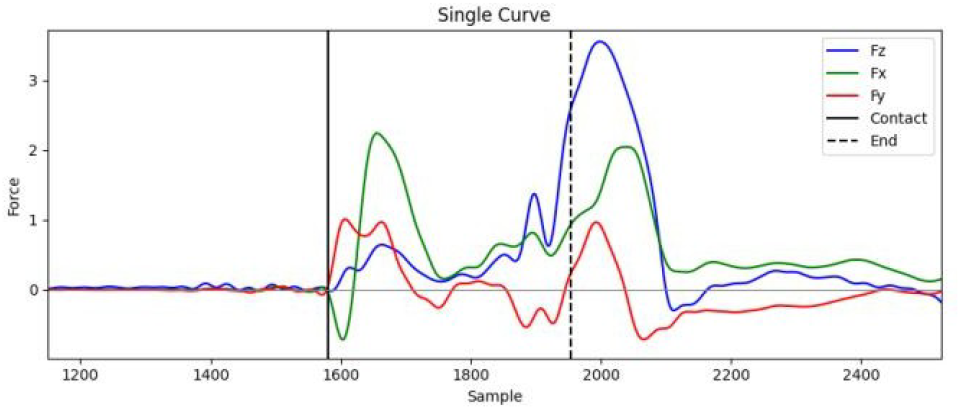
Raw force–time trace for the single-curve wire condition.

**Fig. 5.**
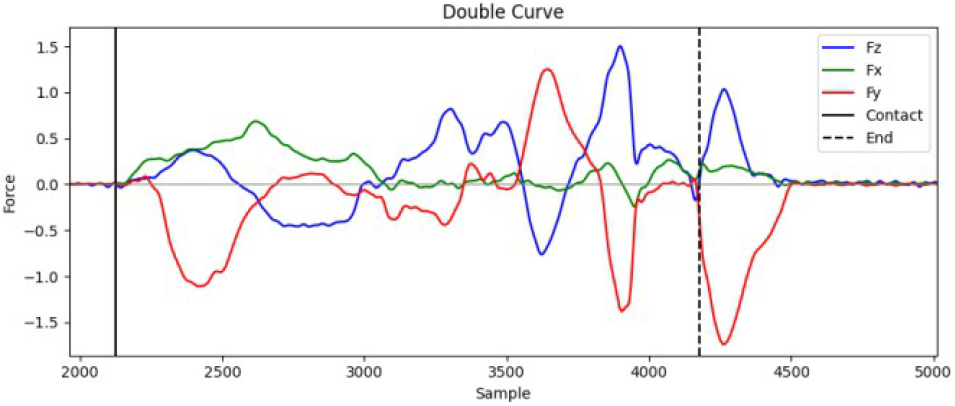
Raw force–time trace for the double-curve wire condition.

### C. Composite Variables

Our goal was to examine different types of variables computed from the BMHFT data. These features were selected to encapsulate important and complementary aspects of motor control, including speed (time), strength (force), smoothness (jerk), and control (impulse). For each force, jerk, and impulse variable, the total of absolute values obtained in each axis (L1 norm) was computed.

1. **Total Duration (time):** Total duration was defined as the time difference between the initial contact with the moving bead and the final contact with bead-to-bead contact at the end of the wire. Let t_c_ denote the timestamp for initial contact and t_e_ the timestamp for task completion.

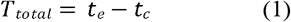
2. **Total Force:** The absolute value of the force components from contact to end was summed to the total force for each trial (the L1 norm). The L1 norm was selected because it analyzes the influence of individual force components through equal weighting and it is more sensitive, allowing for a greater outlier robustness.

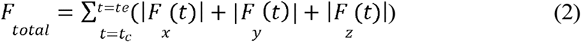
3. **Total Jerk:** The absolute value of the jerk components from contact to end was summed over the duration of the trial. Jerk was defined as the derivative of force with respect to time, and interprets the variability and smoothness of the force application. Higher jerk values indicate inconsistent control and more abrupt motion.

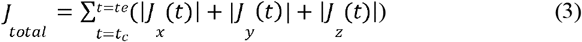
4. **Total Impulse:** Defined as the integral of the magnitude of the force applied across all all axes over the duration of the trial with respect to time. This metric captures total force application while also accounting for task duration, with higher values indicating reduced force control.

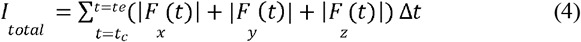

Representative raw force–time traces for each wire condition are shown in **Figs. 3–5**.

### D. Data Preparation

The machine learning analysis utilized a dataset composed of biomechanical features derived from the BMHF device and measurements from the BBT. Extracted variables include total duration (time), total force (TF), total jerk (TJ), total impulse (TI), and Box and Block Test score (BBT). The study included 23 neurotypical participants, with data from both hands from each participant, leading to 46 total samples.

For each participant, there were 5 trials recorded, not including the practice trial. To minimize RMSE and normalize the data for larger external validity, we used a Box-Cox transform and weighted the tails (10th and 90th percentiles). The extracted variables from each of the five trials were averaged, and those values were put in the final datasheet, with the averaged variable data corresponding to the participant.

### E. Machine Learning Analysis

The extracted data set contained multiple biomechanical features, leading to the feature set being higher-dimensional relative to the number of samples. Moreover, multiple of these variables were derived from a common force-time signal, leading to significant multicollinearity. Based on these constraints, we believed that just using standard linear regression could yield inaccurate inference and coefficient estimates with high variance. We used machine learning based approaches that incorporate techniques such as regularization, ensemble learning, and bagging to better identify significant features while also mitigating statistical noise, multicollinearity, and preventing overfitting. The main focus of the models was not to maximize predictive accuracy, but rather to aid in feature ranking based on a correlated feature space.

The regression task aimed to identify significant features for predicting the Box and Block Test score (BBT). Seven machine learning models were chosen to represent key algorithmic categories: Ensemble Methods (XGBoost, Random Forest, Gradient Boost) for their robustness to noise, ability to handle non-linear relationships, and feature importance [21]; Support Vector Regression (SVR) for its effectiveness in high-dimensional spaces via margin maximization and kernel tricks, and Regression Methods (Logistic Regression with L1, L2, and Elastic Net regularization) for their interpretability, sparsity induction and regularization to prevent overfitting in linear models [20,21]. This diverse ensemble ensured a comprehensive assessment of predictive performance, facilitating the identification of optimal models without category bias. Hyperparameter optimization using GridSearchCV with 3-fold stratified cross-validation (StratifiedKFold (n_splits=3, shuffle=True, random_state=42)), enhanced parameter tuning robustness while preserving class distribution. Models were trained and evaluated using a single random seed (42) for reproducibility, reducing computational complexity compared to a multi-seed approach.

The preprocessed dataset was partitioned into training (34 samples) and testing (9 samples) sets using an 80-20 split with random_state=42. Features were standardized using StandardScaler for compatibility with scale-sensitive models like SVR and Logistic Regression. Bagging was incorporated for each model in an effort to increase predictive power. Model performance was evaluated using the test Root Mean Squared Error (RMSE). SHapley Additive exPlanations (SHAP) values were computed for interpretability—using TreeExplainer for ensembles, KernelExplainer for SVR, and LinearExplainer for Logistic Regression—extracting features with non-zero coefficients from both training and test sets [22,23]. The top two models for feature selection were chosen based on the lowest test RMSE.

### F. Feature-Specific Analysis

Computed model-specific SHAP values for the train data were utilized to quantify each feature’s impact on predictions, with features sorted by absolute mean SHAP values, where higher values indicate greater influence on model outputs [22,23]. An Ordinary Least Squares (OLS) Regression was run with the selected variables, and their standardized Beta value was used to determine the relative strength of each predictive variable in relation to the BBT while holding other variables constant. Correlation strengths were interpreted based on standardized Beta values: 0-0.2 (weak), 0.2-0.5 (moderate), 0.5-1 (Strong). The coefficient sign indicates the directional association with BBT.

## III. RESULTS

Out of the seven models chosen, the top two models were the RandomForest and the Logistic Regression (ElasticNet) with test RMSEs of 9.81 and 10.41, respectively (**Fig. 6**). Based on this metric, these models were chosen for further analysis. From these two models, we computed SHAP values to give us information about feature importance. An ordinary multiple linear regression was used to determine the statistical importance of the variable in predicting BBT performance.

**Fig. 6.**
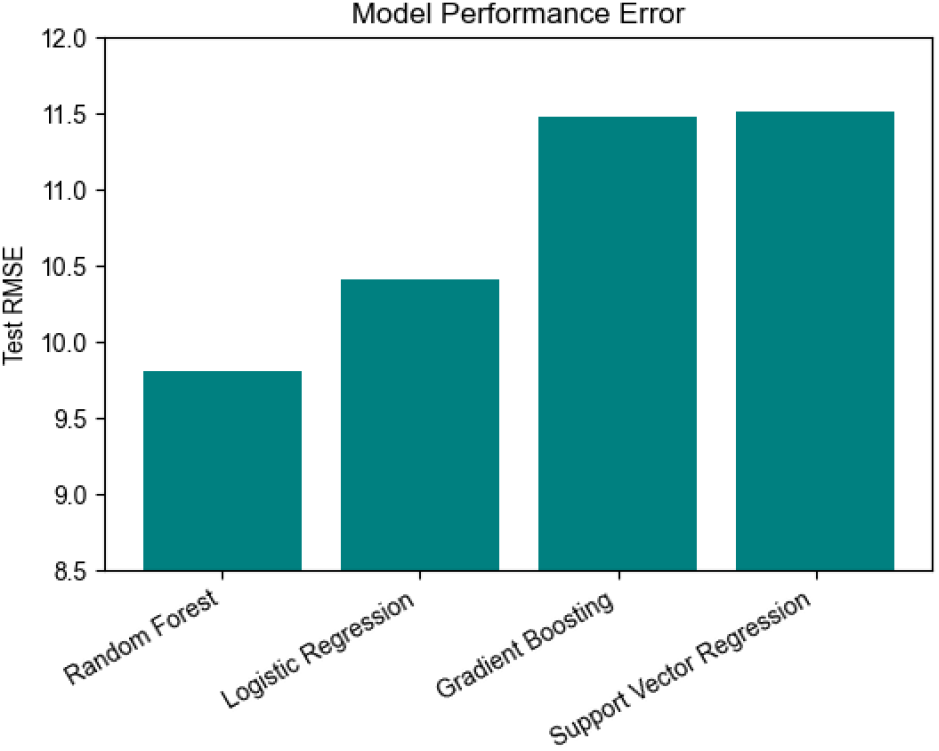
Test root mean squared error (RMSE) for the top four out of the seven total machine learning models. Lower values indicate better performance.

SHAP analysis revealed that task completion time variables contributed most strongly to BBT prediction (**Fig. 7**). Double-curve time and straight wire time showed the highest aggregated mean absolute SHAP values, indicating the greatest influence on model output. This was followed by impulse and jerk variables with comparatively lower contributions. Overall, the SHAP results suggest that temporal aspects of task execution play a more dominant role in predicting BBS than force-based metrics, as hypothesized.

**Fig. 7.**
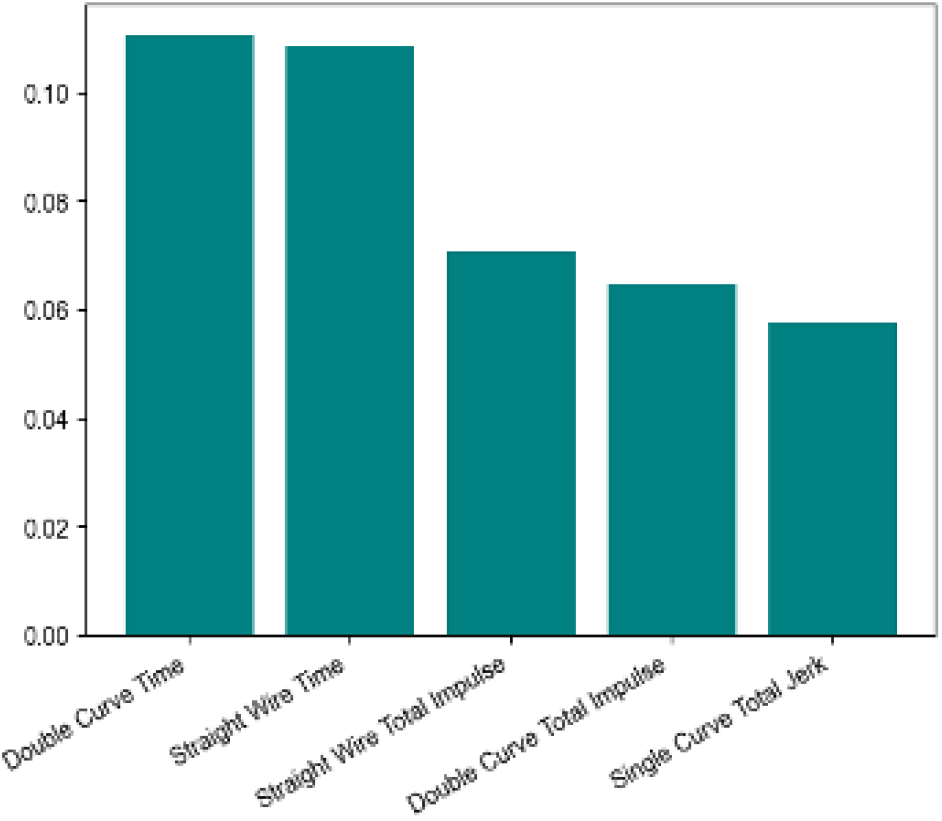
Top five features identified by the Random Forest model, ranked by mean SHAP value.

Multiple linear regression results are summarized in **Table 1**. Among all predictors, double-curve wire time, with the highest SHAP importance, emerged as the only statistically significant variable (β = −1.05, *p* = 0.04), exhibiting a strong negative association with BBT. Other variables, including straight-wire time and impulse-based measures, showed weak to moderate effect sizes but did not reach statistical significance (*p* > 0.05). These findings indicate that while several features contribute to model prediction, only double-curve wire time independently predicts BBT.

**TABLE I.** TOP 5 FEATURES FROM RANDOM FOREST MODEL AND THEIR ASSOCIATION WITH BOX AND BLOCK SCORE.

| Feature | Standardized $\beta$ | Effect Strength | p-value | 95% CI |
| --- | --- | --- | --- | --- |
| Double-Curve Wire- Time | -1.05 | Strong | 0.04 | [-2.04, -0.06] |
| Straight Wire- Time | -0.34 | Moderate | 0.40 | [-1.12, 0.48] |
| Straight wire- total impulse | 0.26 | Weak | 0.45 | [-0.43, 0.96] |
| Double-curve wire-total impulse | 1.13 | Strong | 0.23 | [-0.74, 3.01] |
| Single-curve wire-total jerk | -0.28 | Moderate | 0.1 | [-0.61, 0.06] |

## IV. DISCUSSION

In this study, we analyzed metrics derived from the BMHFT and examined how well they best predicted performance on the Box and Block Test, a common clinical examination used to assess motor function. Our main finding was that total duration (time) on the most complex wire (double curve) was the best predictor of the Box and Block Test score (BBT), whereas force-based predictors were weak predictors of BBT.

The strong correlation between time on the double curve wire and BBT suggests that the duration of an increased complexity task reflects the same motor abilities that are valued in clinical assessments. The BBT primarily emphasizes efficiency during the examination due to its time-constrained nature, with the goal being to move the most blocks in a set period of time. Prior work has shown that this examination mirrors speed-based parts of dexterity rather than fine motor control and execution [18,19]. When the complexity of a motor task increases, there are greater demands regarding anticipation, movement planning, force control, and error minimization [3,4,8]. This makes total duration an important proxy to performance on clinical, time-based measures like the BBT.

On the other hand, the weak correlation between force-based variables and BBT underscores the inability of the BBT to capture the sensorimotor force control behavior. Metrics like total force production (magnitude), variability, smoothness, and control reflect larger sensorimotor processes involved in object manipulation. Prior work has demonstrated that sensorimotor and kinetic metrics may capture aspects of the quality of the movement behavior not found or accounted for in time or score-based clinical exams [9,12]. Therefore, the weak predictive value of force-based variables on BBS should not be interpreted as insignificant, but instead as evidence of clinical examinations like the BBT not accounting for these facets of motor behavior.

This discrepancy between time-based clinical exams and force-based assessments underlines a larger issue in pediatric motor function assessment. Clinicians often design exams in such a way that they are applicable, easy to administer, and efficient. However, this practicality frequently comes at the cost of being unable to quantify changes in movement quality or control strategies, including error control. Moreover, pediatric and developmental research in motor control argues that the combination of sensory, motor, and cognitive systems contributes towards skilled and high-quality behaviors and that movement quality cannot be fully understood based on completion time alone [12].

The findings from this study help support the development and need for a unified metric that links both the efficiency of clinical measures and is attuned to the sensitivity of experimental metrics. The BMHFT incorporates both of those metrics while also varying task complexity, providing a tool to bridge the gap between clinical and laboratory findings while also preventing against a ceiling effect. By developing a unified metric(s), it will improve both clinicians’ and scientists’ ability to study adolescent motor development, allowing for a more accurate assessment of developmental delays and neuro/motor degenerative disorders.

### A. Limitations

The main limitation of this study is the smaller sample size (N=46), specifically preadolescent participants, ages 12–14. So far, the primary focus of the BMHFT has been quantifying performance in the age range of 4–10 years, when force control for manipulating objects is known to develop. Additionally, based on the parallel development of dexterity scores and neuroanatomical development [11,12], we expect that additional participants aged 12–14 years would not significantly change the relationship between total force and age.

### B. Conclusions

We observed how both time-based and force-based variables can predict performance on the BBT, a clinical time-based assessment of hand function. The preliminary evidence shows a strong correlation with time on the double curve wire and BBT, suggesting that duration (time) is an important variable in understanding motor behavior, and that the weak correlation between force-based variables that determine quality of motor behavior and BBT highlights the discrepancy in a holistic assessment of pediatric motor behavior. A better understanding of how the development of motor capabilities and control influences treatment outcomes can aid in the refinement of rehabilitation protocols and treatments [5]. Furthermore, this highlights the necessity of the BMHFT due to its ability to collect multiple types of information related to motor behavior. For clinicians, this could mean a more holistic assessment of a child’s hand function and better-informed intervention planning. The findings suggest further development of sensitive, force-based measures to improve outcomes for children who struggle with fine motor tasks. From this study, future work includes developing a unified score that integrates both time and force-based variables to understand motor control and testing the BMHFT on larger clinical populations.

## Data Availability

All data produced in the present study are available upon reasonable request to the authors

## V. ACKNOWLEDGEMENTS

I acknowledge and thank all the children and their parents for their participation. I would also like to acknowledge the BRAIN program, FDA, and my mentors, Komal K. Kukkar, and Dr. Pranav J. Parikh, for their guidance and support. This project was supported by the UH CLASS RPG (PI: Parikh) and the NIH C-STAR/Shirley Ryan Lab grant (PI: Parikh). Dr. Parikh holds a financial interest in Dextify Innovations LLC. Dextify Innovations LLC would like to develop further and commercialize the Bead Maze device used in this research.

